# Impact of the COVID-19 pandemic on Older Adults Mental Health Services: a mixed methods study

**DOI:** 10.1101/2020.11.14.20231704

**Authors:** Rohan Bhome, Jonathan Huntley, Christian Dalton- Locke, Norha Vera San Juan, Sian Oram, Una Foye, Gill Livingston

## Abstract

**Purpose:** The Covid-19 pandemic is likely to have a significant impact on older adults mental health care. Our study aimed to explore staff perspectives on key challenges and innovations, to help inform the delivery of older adults mental health care in subsequent waves of the pandemic.

**Methods:** A mixed methods online questionnaire developed by NIHR Mental Health Policy Research Unit (MHPRU) was used to gather staff perspectives on their challenges at work, problems faced by service users and their carers, and sources of help and support. Descriptive statistics were used for quantitative analysis and descriptive content analysis for qualitative analysis.

**Results:** 158 participants, working in either community or inpatient settings, and from a range of professional disciplines, were included. For inpatient staff, a significant challenge was infection control. In the community, staff identified a lack of access to physical and social care as well as reduced contact with friends and families as being challenges for patients. Remote working was seen as a positive innovation along with Covid-19 related guidance from various sources and peer support.

**Conclusion:** Our study, with a focus on staff and patient well-being, helps to inform service development for future waves of the pandemic. We discuss measures to improve infection control in inpatient settings, the role of voluntary organisations in supporting socially isolated community patients, the need for better integration of physical and mental health services at an organisational level, and the importance of training staff to support patients and their families with end of life planning.

## Introduction

Older adults are at higher risk of dying from COVID-19 [1], the illness caused by SARS –CoV-2 infection, which was declared a pandemic by the World Health Organisation on 11^th^ March 2020. The UK government, therefore, categorised people aged above 70 as high-risk, regardless of medical co-morbidities, and encouraged them to maintain rigorous social distancing, including not going to other people’s home or outside, with concomitant risks of loneliness [2, 3]. Loneliness can lead to depression and anxiety [4]. Older people [5] and those with severe mental illness or those with underlying chronic physical health illnesses, were more isolated at the beginning of the pandemic and therefore likely to be disproportionally affected [6]. Functional and cognitive impairments experienced by older people with severe enduring mental illness or dementia [7-9] may further exacerbate the impacts of COVID-19 and social isolation. As such, older adults with severe enduring mental illness and people with dementia require special attention from mental health policy makers to ensure robust care provision.

The COVID-19 pandemic presents significant challenges for the delivery of mental health care in general and within older adult services specifically. These challenges come against the backdrop of an already underfunded and under-resourced mental health care system in which older adults are disproportionally disadvantaged [10]. Infection control may be particularly difficult to implement in psychiatric inpatient services because patients are likely to have functional and cognitive impairment which may affect their ability to understand and adhere to infection control guidelines. In the community, challenges may arise managing the needs of vulnerable older patients with little face-to-face contact.

Concerns have been raised about the impacts of the pandemic on the mental health of staff and their wellbeing [11]. Staff have had to cope with redeployment at short notice accompanied by increased risk of contracting the infection and subsequently transmitting it to household members. These risks have been compounded by the need to self-isolate after developing symptoms, inconsistent guidance on the use of personal protective equipment (PPE) and, at times, the inaccessibility of testing [12]. In response to these challenges, UK mental health services have introduced new initiatives and undergone rapid reconfigurations to reduce the risk of infection and the impacts of staff sickness while supporting staff and managing the needs of patients.

Several position pieces have highlighted the potential impact of COVID-19 on mental health services [13, 14]. To our knowledge, a mixed methods study conducted in the UK by the NIHR Mental Health Policy Research Unit (PRU) [15] is the only research to date that captures the views and experiences of people working at the forefront of mental health services during the pandemic. Knowledge of challenges, successful and less useful innovations should, in the short-term, support staff and patient well-being in future waves of COVID-19 and, in the longer-term, inform service development and policymaking for older adults mental health care. Here, we report data from this study on the challenges identified by staff working in older adult mental health services. We also present staff perspectives on which resources and innovations have been beneficial, which should be retained, and which have been difficult to implement.

## Methods

### Participants and procedure

The NIHR Mental Health Policy Research Unit (MHPRU) developed an online questionnaire to collect cross sectional quantitative and qualitative data from mental health care staff working across settings and sub-specialties. They rapidly disseminated the questionnaire through professional networks, social media and relevant mental health-focused bodies, collecting data between 22^nd^ April 2020 and 12 May 2020 [15]. More detailed information on questionnaire design and dissemination is reported in the earlier paper [15].

The present study includes a subset of the participants from that original study, who worked in face-to-face mental health care treating older adult patients or people with dementia. We included all professional groups, such as nurses, psychologists, social workers, peer support workers, occupational therapists and psychiatrists. Participants could work in the NHS, private healthcare, social care or voluntary sector organisations.

To ensure participants were reporting their experience with older adults, we included participants who worked only in older adult inpatient services, community mental health teams (CMHTs) (not providing dementia care) or memory (dementia) services but excluded those who worked with older adult patients as well as another patient group.. To enable comparison between different settings (eg inpatient vs community) we excluded participants who worked in multiple settings.

### Questionnaire content

The questionnaire contained a mixture of structured questions and open-ended questions. Participants were initially asked which sector, setting and mental health speciality they worked in as well as their professional discipline. Additionally, participants were asked which region of the country their service was in and whether it was an area with a population of more or less than 100 000 people.

The core questions of the questionnaire were split into three sections. They were: challenges at work during the COVID-19 pandemic (24 items), perceived problems currently faced by mental health service users and family carers (23 items) and sources of help and support at work during the pandemic (14 items). All participants were asked to rate each item on a five-point Likert scale ranging from ‘not relevant’ to ‘extremely relevant’ for the first two sections and from ‘not at all important to ‘extremely important’ in the third.

The questionnaire also contained a series of open-ended questions. To address our study aims we included questions that explored innovations or initiatives that had worked well, any helpful resources or guidance on managing the impact of the pandemic, any innovations that staff would want to remain in place and any innovations or guidance that were difficult to implement.

There were additional sections of the survey only open to staff working in particular settings or specialties. Of relevance to our study were three sections for staff working in inpatient services, community services and older adult services. Some of the specific items for those working in older adults’ services related to supporting clients who were self-isolating, did not have the usual level of family support, were in residential homes where COVID-19 may have been present, and may have cognitive or sensory impairment. Other items in this section considered end of life planning and reduced community services.

A copy of the survey is available at: https://opinio.ucl.ac.uk/s?s=67819

### Analysis

Quantitative data: Descriptive statistics were produced using Stata 15 to summarise demographic information and participant characteristics such as their professional background, speciality and work setting. Items eliciting staff views were answered on a five-point Likert scale which ranged from ‘not relevant’ to ‘extremely relevant’, except for the ‘sources of help’ section where responses were rated as ‘not important at all’ to ‘extremely important’. Percentages of each response were calculated.

Qualitative data: We carried out qualitative analysis to identify innovations that helped tackle some of the challenges that staff had highlighted in the quantitative analysis. RB identified the main themes about innovations emerging from participants’ open-ended responses and developed a preliminary analytic coding framework based on the study’s aims. Coding matrices were developed using Microsoft Excel, with the emerging codes in columns and participants’ responses in rows. Participant responses to open-ended questions were left unedited and indexed by RB in the matrix under the relevant theme. Descriptive content analysis was conducted [16]. New codes were developed for topics that arose repeatedly but did not fit into the initial coding framework. Coding was discussed with GL, NVSJ, CDL, and JH, who met to refine the emerging codes to ensure all the relevant themes emerging from the data were captured.

## Results

1194 survey participants provided mental health care to older adults of whom 298 (25%) worked only with older adults. Of these 298 participants, 218 (73%) answered at least one question from each of the three core sections of the study. 60 of these participants worked across inpatient and community services and so were excluded. We therefore include 158 participants in our final analyses, 67 (42%) from inpatient settings, 58 (37%) from older adult community mental health teams and 33 (21%) from memory services.

### Participant characteristics

The majority of participants, 142 (90%) out of 157 (1 missing value), were working in their normal setting while 12 (8%) had been redeployed and three (2%) were locum staff. 81% of participants were female (128 of the 158 participants specified their gender). 113 (93%) out of 121 participants (37 missing values) stated their ethnicity as white. Sixty-five (41%) of the participants were nurses, 21 (13%) occupational therapists, 19 (12%), psychologists, 18 (11%) psychiatrists, 8 (5%) peer support workers and one social worker, as well as 26 (18%) who stated their profession or role as ‘other’. Forty-nine (31%) participants had either a managerial or lead clinician role. The vast majority of participants were based in England (132, 84%) while 17 (11%) were in Scotland and 9 (6%) were in Wales. Most participants (109, 69%) were based in cities or towns with populations of more than 100 000 people. Further data on demographics, personal caring responsibilities and COVID-19 status can be found in **supplementary table 1**.

**TABLE 1.**
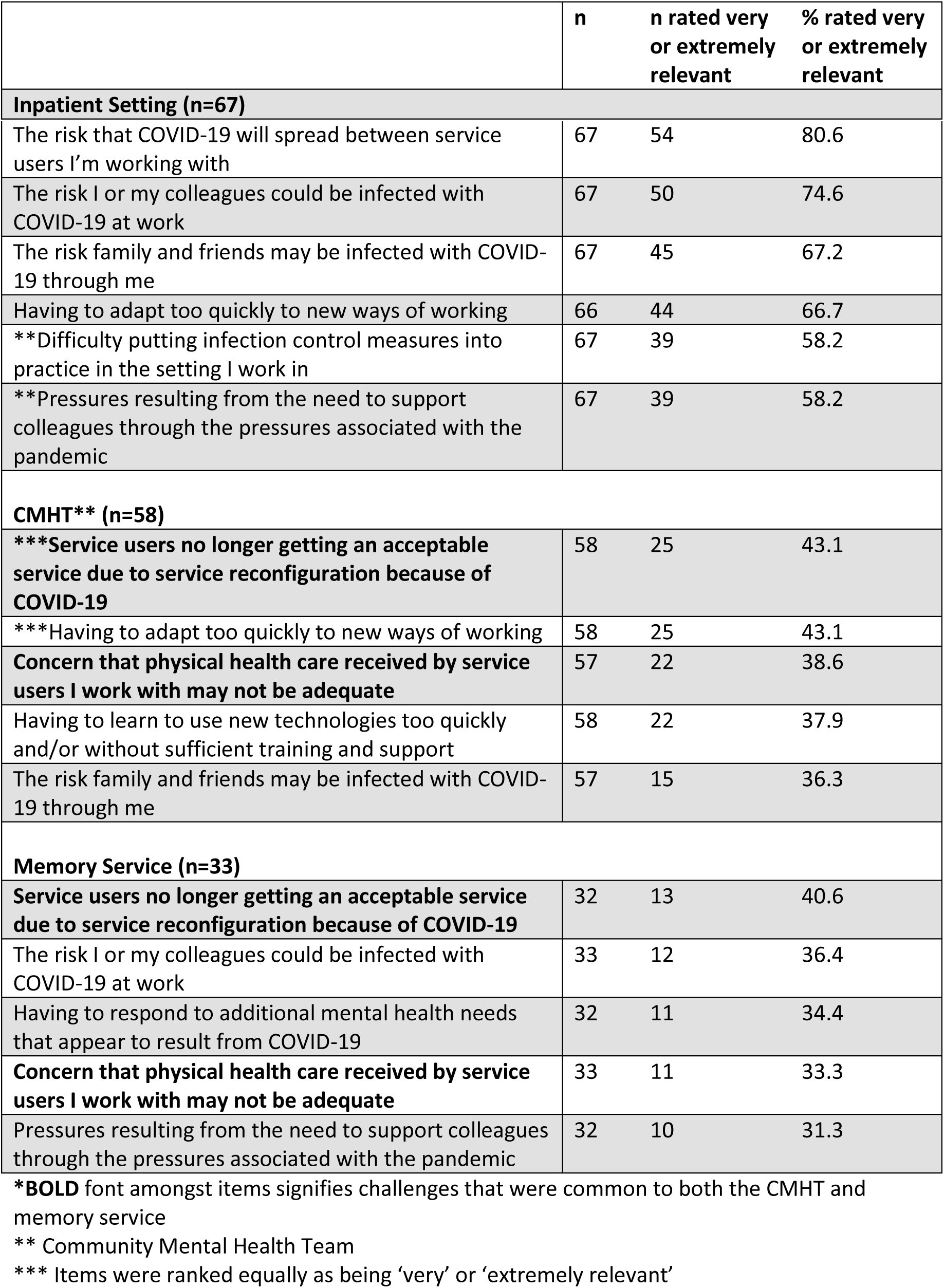
**The top five rated work challenges in each setting, in order of % rated very or extremely relevant**

|  | n | n rated very or extremely relevant | % rated very or extremely relevant |
| --- | --- | --- | --- |
| **Inpatient Setting (n=67)** |  |  |  |
| The risk that COVID-19 will spread between service users I'm working with | 67 | 54 | 80.6 |
| The risk I or my colleagues could be infected with COVID-19 at work | 67 | 50 | 74.6 |
| The risk family and friends may be infected with COVID-19 through me | 67 | 45 | 67.2 |
| Having to adapt too quickly to new ways of working | 66 | 44 | 66.7 |
| **\*\*Difficulty putting infection control measures into practice in the setting I work in** | 67 | 39 | 58.2 |
| **\*\*Pressures resulting from the need to support colleagues through the pressures associated with the pandemic** | 67 | 39 | 58.2 |
| **CMHT\*\* (n=58)** |  |  |  |
| **\*\*\*Service users no longer getting an acceptable service due to service reconfiguration because of COVID-19** | 58 | 25 | 43.1 |
| **\*\*\*Having to adapt too quickly to new ways of working** | 58 | 25 | 43.1 |
| **Concern that physical health care received by service users I work with may not be adequate** | 57 | 22 | 38.6 |
| Having to learn to use new technologies too quickly and/or without sufficient training and support | 58 | 22 | 37.9 |
| The risk family and friends may be infected with COVID-19 through me | 57 | 15 | 36.3 |
| **Memory Service (n=33)** |  |  |  |
| **Service users no longer getting an acceptable service due to service reconfiguration because of COVID-19** | 32 | 13 | 40.6 |
| The risk I or my colleagues could be infected with COVID-19 at work | 33 | 12 | 36.4 |
| Having to respond to additional mental health needs that appear to result from COVID-19 | 32 | 11 | 34.4 |
| **Concern that physical health care received by service users I work with may not be adequate** | 33 | 11 | 33.3 |
| Pressures resulting from the need to support colleagues through the pressures associated with the pandemic | 32 | 10 | 31.3 |

\***BOLD** font amongst items signifies challenges that were common to both the CMHT and memory service
\*\* Community Mental Health Team
\*\*\* Items were ranked equally as being 'very' or 'extremely relevant'

### Challenges at work during the COVID-19 pandemic

**Table 1** shows the five highest rated work challenges in each setting out of the possible 24. In inpatient settings, key challenges centred around infection control, with staff reporting concerns about transmission between patients and to staff and about the risk of staff transmitting the infection to family and friends. Adapting to new ways of working and supporting colleagues under pressure were also highlighted as challenges.

In community settings, across both CMHT and memory services, staff were concerned that the patients that they cared for may not receive adequate physical healthcare service and that service reconfigurations secondary to COVID-19 may lead to suboptimal mental health care. In community mental health teams, additional challenges identified were the risk of respondents transmitting COVID-19 to family and friends and having to adapt to new ways of working, including having to learn to use new technologies without adequate support. In memory service settings, staff also reported the risk of being infected by the virus at work and pressures associated with supporting colleagues with COVID-19 related concerns as being important challenges. Participants who responded to the section of the survey specifically designed for those working in community settings, highlighted the challenge of providing sufficient support with reduced staffing and face-to-face contact (**supplementary table 2**).

**Table 2.** **Staff perspective on patients’ and carers’ problems that were most relevant during the COVID-19 Pandemic, in order of % rated very or extremely relevant**

| **Inpatient Setting (n=67)** | **n** | **n rated very or extremely relevant** | **% rated very or extremely relevant** |
| --- | --- | --- | --- |
| Lack of access to usual support networks of friends and family | 67 | 55 | 82.1 |
| Worries about family getting COVID-19 infection | 67 | 49 | 73.1 |
| **\*\*Worries about getting COVID-19 infection** | 67 | 44 | 65.7 |
| **\*\*Loneliness due or made worse by social distancing, self-isolation and/or shielding** | 67 | 44 | 65.7 |
| High personal risk of severe consequences of COVID-19 infection (eg. due to physical health comorbidities) | 67 | 43 | 64.2 |
| **CMHT\*\*\* (n=58)** |  |  |  |
| **Loneliness due or made worse by social distancing, self-isolation and/or shielding** | 58 | 50 | 86.2 |
| **Lack of access to usual support networks of friends and family** | 58 | 49 | 84.5 |
| **Lack of access to usual support from other services (primary care, social care, voluntary sector)** | 58 | 46 | 79.3 |
| Increased difficulties for families/carers | 57 | 44 | 77.2 |
| Lack of usual work and activities | 57 | 42 | 73.7 |
| **Memory Service (n=33)** |  |  |  |
| **Lack of access to usual support networks of friends and family** | 33 | 29 | 87.9 |
| **Loneliness due or made worse by social distancing, self-isolation and/or shielding** | 33 | 26 | 78.8 |
| **Lack of access to usual support from other services (primary care, social care, voluntary sector)** | 33 | 22 | 67.7 |
| High personal risk of severe consequences of COVID-19 infection (eg. due to physical health comorbidities) | 32 | 20 | 62.5 |
| **\*\*Worries about family getting COVID-19 infection** | 33 | 20 | 60.6 |
| **\*\*Lack of usual work and activities** | 33 | 20 | 60.6 |

\***BOLD** font signifies challenges that were common to both the CMHT and memory service
\*\*Items were ranked equally as being 'very' or 'extremely relevant'
\*\*\*Community Mental Health Team

### Staff perspectives on difficulties faced by patients and carers during the COVID-19 pandemic

**Table 2** summarises staff perspectives on the key problems for patients and carers that they are in contact with. Throughout all settings, staff rated the relevance of loneliness due to social distancing measures and lacking access to usual support networks very highly. Inpatient and memory service staff were concerned about the risk of severe consequences of COVID-19 infection amongst their patients. In inpatient settings, staff also thought that patients’ concerns included them or their family members getting infected with COVID-19. In both community settings, the loss of usual support from primary care, social services and voluntary sector organisations was seen as a difficulty for patients. In CMHT settings, the lack of usual work and activities and stress on families and carers were judged to be other areas of concern.

### Sources of help and support for staff in the workplace during the COVID-19 pandemic

**Table 3** summarises the most relevant sources of help and support for staff working during the COVID-19 pandemic. Throughout all settings, support and information from colleagues as well as guidance at a local (employer) and national (NHS, professional bodies) level were regarded as most helpful. Inpatient staff also found support from managers and the wider public support for keyworkers to be helpful. In community settings, staff thought that the resilience and resourcefulness of patients and carers were important. In CMHTs, staff thought the adoption of new digital ways of working were beneficial while in the memory service support from voluntary sector organisations was recognised as being helpful.

**Table 3.**
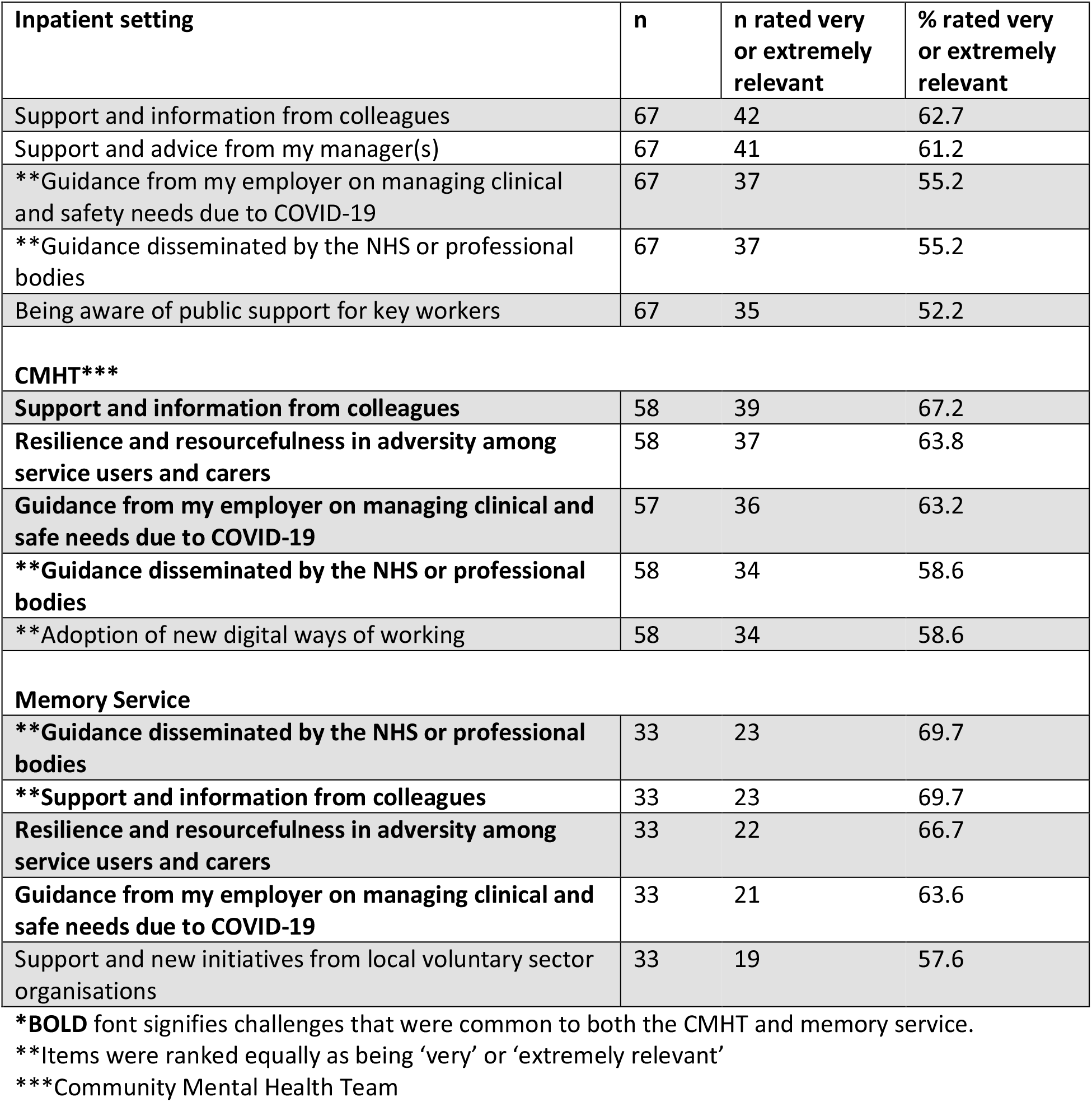
**Top five sources of help and support for staff, in order of % rated very or extremely relevant**

| **Inpatient setting** | **n** | **n rated very or extremely relevant** | **% rated very or extremely relevant** |
| --- | --- | --- | --- |
| Support and information from colleagues | 67 | 42 | 62.7 |
| Support and advice from my manager(s) | 67 | 41 | 61.2 |
| **\*\*Guidance from my employer on managing clinical and safety needs due to COVID-19** | 67 | 37 | 55.2 |
| **\*\*Guidance disseminated by the NHS or professional bodies** | 67 | 37 | 55.2 |
| Being aware of public support for key workers | 67 | 35 | 52.2 |
| **CMHT\*\*\*** |  |  |  |
| **Support and information from colleagues** | 58 | 39 | 67.2 |
| **Resilience and resourcefulness in adversity among service users and carers** | 58 | 37 | 63.8 |
| **Guidance from my employer on managing clinical and safe needs due to COVID-19** | 57 | 36 | 63.2 |
| **\*\*Guidance disseminated by the NHS or professional bodies** | 58 | 34 | 58.6 |
| **\*\*Adoption of new digital ways of working** | 58 | 34 | 58.6 |
| **Memory Service** |  |  |  |
| **\*\*Guidance disseminated by the NHS or professional bodies** | 33 | 23 | 69.7 |
| **\*\*Support and information from colleagues** | 33 | 23 | 69.7 |
| **Resilience and resourcefulness in adversity among service users and carers** | 33 | 22 | 66.7 |
| **Guidance from my employer on managing clinical and safe needs due to COVID-19** | 33 | 21 | 63.6 |
| Support and new initiatives from local voluntary sector organisations | 33 | 19 | 57.6 |

**\*BOLD** font signifies challenges that were common to both the CMHT and memory service.
**\*\***Items were ranked equally as being 'very' or 'extremely relevant'
**\*\*\***Community Mental Health Team

In the section of the survey completed only by staff working in older adults services (**supplementary table 3**), 62% (31/50) of staff working in inpatient services thought that increased involvement in end of life planning was very or extremely relevant, while the number was far lower in CMHT (20%, n=40) and memory service (18.18%, n=33) settings.

### Quantitative analysis

#### Innovation and resources that staff found helpful

**Table 4** summarises the analyses of innovations and resources that staff reported as having been helpful. Remote working was identified as being helpful across all settings. For inpatients, it enabled greater attendance of ward rounds by multidisciplinary professionals involved in an individual’s care. Staff working in the community reported that not travelling to appointments was more time efficient and remote working provided an opportunity for those in the community to work from home, which helped to reduce the risk of transmitting or acquiring COVID-19. While generally construed as being a positive innovation, staff suggested that it limited peer support.

**Table 4.** **Innovations and resources that staff found helpful during the COVID-19 pandemic**

| **Innovations** |  |
| --- | --- |
| **Inpatients** | **Remote working**<br>Virtual multidisciplinary team meeting for patients facilitating broader attendance<br>**Staff**<br>Flexibility in working patterns<br>**Patients**<br>Facilitating contact with family and friends using video calls |
| **CMHT\*** | **Remote working**<br>Reduced travelling time which allows more patients to be assessed.<br>Opportunity to work from home<br>Patients in rural areas could be accessed more readily<br>**Staff**<br>Flexibility in working patterns |
| **Memory Service** | **Remote working**<br>More time efficient<br>Fewer "Did not attend" (missed appointment)<br>Opportunity to work from home<br>**For Staff**<br>Flexibility in working patterns<br>Staff uniforms<br>**For Patients**<br>Well-being phone calls<br>Risk stratification to identify most vulnerable patients |
| **Resources** |  |
| **Inpatients** | **For staff**<br>Staff meetings to discuss concerns<br>**Guidance**<br>From professional bodies, eg. British Psychological Society, Royal College of Occupational Therapists<br>Local (trust), national (Public Health England) and international (World Health Organisation) |
| **CMHT\*** | **For staff**<br>Informal peer support<br>Psychology led reflective groups<br>**For patients**<br>Wellbeing packs<br>Voluntary organisations providing practical support<br>**Guidance**<br>Local (daily updates from the trust), national (Alzheimer's Society, British Geriatrics Society)<br>Posters<br>Personal protective equipment |
| **Memory Service** | **For staff**<br>Mindfulness sessions<br>Informal peer support<br>Staff helpline<br>**For patients**<br>Activity packs for patients<br>Voluntary organisations providing practical support, eg shopping<br>**Guidance**<br>Local (chief executive daily update, intranet), national (Alzheimer's Society)<br>Webinars<br>Specialist guidelines, eg end of life care<br>Personal protective equipment |

\*Community Mental Health Team

> ‘*Home based working has been effective in supporting staff to reduce anxieties and engage with their caseloads remotely whilst minimising risk of exposure to themselves, their families and the patients within their caseloads*.’ (Occupational Therapist, memory service)

The use of technology to facilitate remote working and patient assessments appears to have been well received across all settings.

> *‘Immediate and widespread use of virtual meetings and consultations. Telephone consultations working particularly (and surprisingly) well with our older adult patient group and their carers. Use of IT to facilitate long-distance ward rounds to reduce infection and travel has tremendous promise*.*’* (Psychiatrist, inpatients)

Across all settings, staff highlighted that the increased flexibility in working was helpful. They also felt that a variety of ways of making increased peer support available, including through psychology led reflective groups and telephone helplines, was a positive intervention in helping to manage the psychological impact of COVID-19 on staff.

> ‘*Coping with Covid-19 staff support helpline manned by psychology staff. Staff support consultation sessions: mindfulness, moral injury and coping strategies. Staff Facebook page to share resources to support staff. Microsoft Teams. Staff Carers forum webinar for staff who are also carers for loved ones. Looking after yourself resource on the intranet*.’ (Clinical Psychologist, memory service)

In community services, staff highlighted patient ‘well-being’ packs and practical support provided by voluntary services for patients as being important resources for patients.

> ‘*Local voluntary groups are helping to provide support for shopping*.*’* (Occupational Therapist, memory service).

Staff in memory services highlighted well-being phone calls and risk stratification of caseloads to identify those who were most vulnerable as positive innovations in response to COVID-19.

The majority of participants thought that the guidance issued at local, national and international level was helpful. In particular, several participants highlighted that guidance from their professional body targeted towards their professional role was beneficial.

> ‘*RCOT [Royal College of Occupational Therapists] guidance on social distancing and covid rehabilitation expectations. OT [Occupational Therapy] guidance from Australia and Illinois university addressing impact of COVID on occupational participation and engagement during and post covid*.*’* (Occupational Therapist, memory service)

#### Innovations that staff would want to remain in place

**Table 5** shows some innovations and changes that staff would like to remain in place. Across all settings, the use of technology to facilitate remote communication and working were frequently highlighted as efficient and sometimes leading to better communication with patients and families.

**Table 5.** **Innovations and changes that staff would like to remain in place and challenges associated with implementing certain innovations**.

| **Innovations to remain in place** |  |
| --- | --- |
| **Inpatients** | **For patients**<br>Facilitating contact with family and friends with video calls<br>Improved focus on physical health<br>**For staff**<br>Easier access to parking<br>Free meals<br>Better forums to discuss concerns with colleagues |
| **CMHT\*** | **For patients**<br>Well-being phone calls<br>More out of hour cover for services<br>**For staff**<br>Virtual team meetings |
| **Memory Service** | **For staff**<br>Frontline workers being involved in management decisions<br>Flexibility in working patterns |
| **Challenges with implementing certain innovations and guidelines** |  |
| **Inpatients** | Guidelines changing frequently<br>Contradictory guidelines<br>Difficulty in implementing social distancing in this patient group<br>Use of personal protective equipment |
| **CMHT\*** | Hierarchical dissemination of information<br>Lack of testing<br>No clear guidance on what is considered urgent or severe enough to warrant a face-to-face home visit |
| **Memory Service** | Working from home does not allow same level of peer support<br>Some patients cannot use the technology for video calls<br>Some guidelines are developed without an understanding of the practicalities |

\*Community Mental Health Team

> ‘*Keep ward mobile phone for patient use. Option for relatives to phone in for CPA meetings (if unable to attend face to face) after pandemic’*. (Occupational Therapist, inpatients).

> *‘I would like to continue to have team meetings via videocall where these are not at my usual base as otherwise this involves a significant loss of working time’*. (Clinical Psychologist, CMHT)

Additionally, inpatient staff suggested that the improved focus on physical health should remain in place to benefit patients in future. Some inpatient staff also suggested that free meals and easier access to parking should continue for staff.

In the CMHT, a few participants thought that greater out of hours service provision should remain in place. In the memory service, staff suggested that they would like to see frontline mental health workers continuing to contribute to management decisions.

#### Innovations or guidance that staff found difficult to implement

**Table 5** also highlights innovations or guidance that staff found difficult to implement. This does not necessarily mean that the innovation was ineffective or unhelpful but certain factors made it difficult to put into practice. The use of technology to enable remote patient contact was previously highlighted as being a beneficial innovation. However, some staff in the CMHT felt that there needed to be clearer guidance on which patients required a face-to-face assessment. Further, in both CMHT and memory services, respondents stated that some of their patients could not utilise the technology required for remote assessments effectively.

> *‘I work with older people and many are unable to use technology’*. (Nurse, CMHT)

Social distancing, while clearly an important infection control measure, was difficult to implement in inpatient settings.

> ‘*Working with patients with moderate-advanced dementia who are unable to understand about the coronavirus, therefore unable to follow restrictions/ social distancing. Unable to social distance in offices due to reduced space/number of staff*’. (Occupational Therapist, inpatients)

Although guidance from various sources was seen as helpful, some inpatient staff found it difficult that guidelines changed frequently and could be contradictory. While, in the community, staff thought that guidelines may be developed without a clear understanding of the practicalities and the dissemination could be quite hierarchical.

Staff in community teams thought that the personal protective equipment (PPE) guidance was helpful but the lack of COVID-19 testing was highlighted as being challenging. Only 39% of inpatient staff (n=57) thought that the lack of PPE was very or extremely relevant (**supplementary table 2**). However, in the qualitative analysis, for inpatient staff, the lack of PPE was highlighted as a barrier to infection control. Further, some inpatient staff found the PPE guidance difficult to interpret or implement.

> *‘No PPES* [Sic] *and no facility to wash ourselves or clothes at work. We are forced to take the infection home and then clean it’*. (Other worker, inpatients).
>
> *‘PPE guidelines appear to be interpreted in different ways by different teams’*. (Clinical Psychologist, inpatients).

## Discussion

To our knowledge, this is the first study of the experience and views of staff, who worked in older adult mental health services, in relation to care provision during the beginning of the COVID-19 pandemic. We found that the key challenges for inpatient staff surrounded controlling the transmission of COVID-19. In the community, important challenges were lack of access for patients to usual services for their physical health or social care and to their family and friends. Remote working, guidance from a variety of sources and peer support were seen as being helpful.

There are several similarities between the experiences of staff working in older adult settings and those of staff working across the range of mental health services [15]. Infection control in inpatient settings was seen as a significant concern while remote working was positively received. Staff working in older adult community settings had greater concern about the physical healthcare that their patients would have access to and their patients’ abilities to use technology compared to staff working throughout all mental health settings.

The challenges surrounding infection control in older adult inpatient mental health settings are significant and borne out by a recent study [17] which found that around 40% of patients in a cohort of 131 patients are likely to have contracted COVID-19 whilst an inpatient. The study, which analysed data that was collected at the beginning of the pandemic (1^st^ March 2020 to 30^th^ April 2020) suggested that the lack of testing for infection, poor availability of PPE, asymptomatic carriers and false negative tests contributed to the high infection rate. Looking ahead, access to testing and PPE as well as self-isolation for two weeks of all new patients on the ward will be important in addressing the challenges surrounding infection control. Further, if they continue to be used then staff will have to keep in mind the low sensitivity of polymerase chain reaction (PCR) testing and use clinical judgement to guide their management of symptomatic patients who may have false negative tests.

Staff perceived that a lack of access to their usual support networks and loneliness would be a significant challenge for older adults with mental illnesses or dementia. Accordingly, support provided by voluntary service organisations and well-being packs produced for patients were seen as helpful innovations during the pandemic. Charitable and voluntary organisations, such as *Age UK* and the *Alzheimer’s society*, provided practical and emotional support including help with shopping and telephone calls to reduce loneliness, during the pandemic. Mental health teams, social care services and voluntary organisations should liaise closely to ensure that support is delivered in an organised way, not duplicated and that patients are not overlooked. Social prescribing was also facilitated by online technologies with older adults being supported to access online games, concerts and religious services [18]. However, some older adults are not able access online technologies because they lack equipment, skills, or language proficiency. For these older adults, well-being packs, which could include educational information, sources of support and activities, may be particularly helpful, as well as liaising with their families to help support them. To improve the patient experience, facets of the well-being pack could be individualised in the future.

In our study, staff were concerned that patients in community settings may not be able to or be willing to access physical healthcare services. Reasons for this have been highlighted elsewhere, including the unintended consequences of social distancing messages and strategies aimed at reducing COVID-19 transmission [19]. There has been an emphasis on improved integration of physical and mental healthcare for some time [20]. While it is unrealistic to overhaul the structure of healthcare trusts, increased integration of physical and mental healthcare services at a local level is needed. For example, Tracy et al. [21] reported that older adult inpatient mental health wards being supported by a respiratory nurse specialist and consultant geriatrician was seen as being beneficial during the pandemic. In the community, there need to be clear and agreed pathways for how vulnerable older adults, and especially those with severe and enduring mental illness, can access physical healthcare.

Our study found that remote working has been positively received by staff working in older adult mental health services. Participants reported that remote working has enabled greater flexibility in working patterns, improved time efficiency and allowed the participation of a broader range of professionals in multidisciplinary team (MDT) meetings. These experiences have been reported across mental health services globally [22] and seem likely to be retained once the pandemic abates. However, staff in memory services note that some patients were unable to use technologies that enable remote contact with staff. Some patients, including those unable to access or use technology for virtual assessments, with more complex needs or poor engagement, need face-to-face assessment. Qualitative analyses highlighted the need for clarity on which patients should be offered face-to-face appointments. Presumably, these decisions need to be made on an individual basis based on risk and need.

Guidance issued at local, national and international levels was reported as being helpful by staff. In subsequent waves of the pandemic, it is important that guidance continues to be produced and disseminated in a timely manner. Some inpatient staff commented that guidance changed frequently and in some cases was conflicting. While this is relevant, it is inevitable in a new illness with rapidly evolving knowledge. It might be useful if guidance states that it is based on the best current knowledge but will change as more is learned.

Interestingly, the majority of staff working in inpatient settings thought that their involvement in end of life planning was highly relevant whereas the same applied for only about a fifth of staff working in the community. This may be because in inpatient wards mental health staff give most of the physical care and make decisions about transfer for escalation of physical healthcare. This is not the case in the community but there is an increased risk of death from COVID-19 among older adults [1]. Such planning could provide support to patients’ relatives too given that patients with COVID-19 may experience rapid deterioration and there are often difficult and stressful decisions to be made about whether the ill person should be hospitalised [23]. The relevance of end of life planning, particularly for inpatient staff in this study, highlights the need for appropriate training to enable staff to facilitate discussions about end of life care [24].

### Limitations

Our study has several limitations. There is a risk of sampling bias given that the survey was disseminated through channels which may not have been accessed by all mental health staff. Further, respondents may overly represent those who had strong feelings about the impact of the COVID-19 pandemic and therefore wished to have a platform to voice these. As in the broader survey, non-white mental health staff would appear to be underrepresented [15].

Our study sought to consider the challenges faced by older adult mental health services and the implications for the future, especially in the context of subsequent waves of infection in the pandemic. To truly do this, it would be important to gather perspectives from patients and carers too. While our work did consider patients’ and carers’ difficulties during the pandemic, these were from the perspective of staff.

We were unable to evaluate the impact of COVID-19 on the delivery of older adult mental health care in care homes, due to the limited number of these respondents. This forms an important group of patients especially given the significant mortality and challenges faced by care homes during this pandemic [25].

### Implications

In inpatient settings, clear protocols for infection control and access to appropriate PPE will be important in subsequent waves of COVID-19. In the community, the impact of the loss of patients’ usual support networks may be mitigated through the help provided by third sector organsiations, as well as remote care from statutory services. To facilitate this, there needs to be close liaison between mental health, social care and voluntary services. At an organisational level improved integration of physical and mental health services may greatly benefit metal health patients. Finally, a greater emphasis on training staff to help patients and families in end of life decisions may help patients have a better end-of-life given the high risk of mortality from COVID-19 among older patients.

Future research should seek patients and carer’s perspectives on the impact of the pandemic on mental health services received, including that delivered in care homes.

## Data Availability

The survey dataset is currently being used for additional research by the author research group and is, therefore, not currently available in a data repository. A copy of the survey is available at this web address: https://opinio.ucl.ac.uk/s?s=67819.

## Declarations

### Ethics approval (include appropriate approvals or waivers)

The King’s College London research ethics committee approved the original study (MRA-19/20-18372). Our work is further analysis of data collected during the original study.

### Consent to participate

Information on participation was provided on the front page of the survey. By starting the survey, participants agreed that they had read and understood all this information.

### Consent for publication (include appropriate statements)

It was explained on the front page of the survey that responses may be used in articles published in scientific journals and that these articles will not include any information which could be used to identify any participant.

### Code availability

Not applicable

## Authors’ contributions

The study was conceived by GL, JH and CDL. The quantitative data analysis plan was developed by CDL and conducted by RB. NVSJ planned the qualitative analysis which was conducted by RB, with contributions from NVSJ, CDL, JH and GL. RB drafted the paper and all authors contributed to and approved the final manuscript.

