## supplementary table for "Impact of the COVID-19 pandemic on Older Adults Mental Health Services: a mixed methods study"

**Supplementary tables**

**Supplementary Table 1: Participant demographics (n=158)**

| | **Inpatients (n=67) (%)** | **CMHT* (n=58) (%)** | **Memory service (n=33) (%)** |
|---|---|---|---|
| **Gender** | **(n=55)** | **(n=40)** | **(n=33)** |
| Female | 44 (80) | 34 (85) | 27 (81.8) |
| Male | 11 (20) | 6 (15) | 6 (18.2) |
| **Ethnicity** | **(n=51)** | **(n= 40)** | **(n=30)** |
| White | 48 (94.1) | 37 (92.5) | 28 (93.3) |
| Asian | 2 (3.9) | 1 (2.5) | 0 (0) |
| Black | 1 (2) | 2 (5) | 0 (0) |
| Mixed | 0 (0) | 0 (0) | 2 (6.7) |
| **Profession** | | | |
| Clinical psychologist | 4 (6) | 10 (17.2) | 5 (15.2) |
| Nurse | 28 (41.8) | 22 (37.9) | 15 (45.5) |
| Occupational therapist | 7 (10.4) | 10 (17.2) | 4 (12.1) |
| Peer support worker | 6 (9) | 2 (3.4) | 0 (0) |
| Psychiatrist | 3 (4.4) | 10 (17.2) | 5 (15.2) |
| Social worker | 0 (0) | 0 (0) | 1 (3) |
| Other | 19 (28.4) | 4 (6.9) | 3 (9.1) |
| **Participant working in usual work setting?** | | **(n=57)** | |
| Yes | 54 (80.6) | 56 (98.2) | 32 (97) |
| No, locum | 2 (3) | 1 (1.8) | 0 (0) |
| No, redeployed | 11 (16.4) | 0 (0) | 1 (3) |
| **Manager or lead clinician** | 17 (25.3) | 19 (32.8) | 13 (39.4) |
| **Country** | | | |
| England | 55 (82.1) | 49 (84.5) | 28 (84.8) |
| Scotland | 7 (10.4) | 5 (8.6) | 5 (15.2) |
| Wales | 5 (7.5) | 4 (6.9) | 0 (0) |
| **City or town with population >100 000** | 46 (68.7) | **(n=57)** 43 (75.4) | 20 (60.6) |
| **Caring Responsibilities** | **(n=55)** | **(n=40)** | **(n=33)** |
| For children | 17 (30.1) | 16 (40) | 15 (45.5) |
| For elderly, relatives or friends | 16 (29.1) | 12 (30) | 7 (21.2) |
| **Covid status** | **(n=55)** | **(n=40)** | **(n=32)** |
| Yes, confirmed | 1 (1.8) | 0 (0) | 0 (0) |
| Yes, suspected | 14 (25.5) | 12 (30) | 7 (21.9) |
| No | 40 (72.7) | 28 (70) | 5 (15.6) |

*CMHT (Community Mental Health Team)

**Supplementary table 2. Challenges related to working in specific settings (non-core questions)**

| Most relevant challenges | n (%)* | Least relevant challenges | n (%)* |
|---|---|---|---|
| **Inpatient** | | | |
| Difficulty maintaining infection control because inpatients/residents are too unwell to follow procedures (n=58) | 43 (75.4) | Difficulty meeting the physical health needs of people with confirmed/suspected COVID-19 on the ward (n=58) | 19 (32.8) |
| Difficulty discharging people because services usually available in community are closed or less available (n=58) | 39 (67.2) | Reduced access to advocacy and appeal processes under the Mental Health Act (n=58) | 22 (37.9) |
| Challenges protecting people at high risk of severe COVID-19 infection adequately (n=58) | 38 (65.5) | Risk to staff because of a lack of PPE (Personal Protective Equipment) (n=57) | 22 (38.6) |
| **CMHT**** | | | |
| **Difficult providing sufficient support with reduced numbers of face to face contacts (n=43)** | 28 (65.1) | **Increased work to engage and support homeless people in the area (n=41)** | 2 (4.8) |
| Challenges supporting clients in residential settings (eg due to infection control challenges) (n=41) | 26 (63.4) | Increased demand to large numbers of referrals (n=43) | 3 (7.1) |
| Not being able to depend on other services that are normally available in the community (n=43) | 26 (61.9) | **Problems ensuring clients have medication (n=43)***** | 9 (20.9) |
| | | Problems ensuring safe continuation of medication that requires administration or monitoring in person, e.g. depots, clozapine, lithium (n=43)*** | 9 (20.9) |
| **Memory Service** | | | |
| Challenges supporting clients in residential settings (eg due to infection control challenges) (n=33) | 18 (54.6) | **Increased work to engage and support homeless people in the area (n=33)** | 0 (0) |
| **Difficult providing sufficient support with reduced numbers of face to face contacts (n=33)** | 17 (51.5) | Greater difficulty than usual in arranging hospital admission (n=33)*** | 4 (12.1) |
| Difficulty assessing clients by phone or video call (n=33) | 16 (48.5) | **Problems ensuring clients have medication (n=33)***** | 4 (12.1) |

*The table shows the three challenges that were most and least frequently rated as very or extremely relevant. These items were in the non-core section of the survey and participants only responded if they worked in that particular setting, eg. Inpatient or community (CMHT or memory service).
**Community mental health team (non-memory)
***Two items that were equally ranked as being ‘very’ or ‘extremely relevant’ in the CMHT and memory service.

| Supplementary table 3. Challenges specifically related to working in older adult services | | | |
|---|---|---|---|
| | **Inpatients (n=50)** | **CMHT* (n=40)** | **Memory service (n=33)** |
| Challenges supporting clients and staff in nursing homes and care homes where there is or may be COVID-19 infection | **(n=49)** 24 (49) | **(n=39)** 30 (76.9) | **(n=32)** 17 (53.1) |
| Difficulty reaching clients and carers who are self-isolating or shielded currently | 16 (32) | 28 (70) | 19 (57.6) |
| Difficulties supporting clients who do not have their usual level of family support | 29 (58) | 32 (80) | 24 (72.7) |
| Difficulty engaging remotely with people with cognitive or sensory impairments | **(n=49)** 23 (46.9) | 34 (85) | 24 (72.7) |
| Increased pressures because of reduced levels of social care, primary care, physical health and other community services supporting older people | 28 (56) | 29 (72.5) | 18 (54.6) |
| Increased need for involvement in end of life planning | 31 (62) | 8 (20) | 6 (18.2) |

Number (%) of participants who rated each item as very or extremely relevant from each setting.
*Community mental health team
